# CURRENT POLICIES AND PRACTICES FOR TESTING AND TREATMENT OF CHRONIC HEPATITIS B AND C INFECTION IN HIGH-BURDEN COUNTRIES IN THE WHO EUROPEAN REGION

**DOI:** 10.64898/2026.01.25.26344684

**Authors:** Machiko Otani, Marcelo Contardo Moscoso Naveira, Stela Bivol

## Abstract

Chronic hepatitis B and hepatitis C remain major public health concerns in the WHO European Region, where an estimated 10.6 million people were living with HBV infection and 7.7 million with HCV infection in 2022. Despite this substantial burden, diagnosis and treatment coverage remain low, posing a significant challenge to achieving the WHO regional goal of viral hepatitis elimination by 2030. To assess country-level progress and gaps in hepatitis testing and treatment, the WHO Regional Office for Europe conducted a survey among nine high burden countries, receiving responses from eight: Armenia, Azerbaijan, Belarus, Kazakhstan, Kyrgyzstan, the Republic of Moldova, Turkmenistan and Uzbekistan. The survey examined national policies, testing availability, treatment regimens, service delivery approaches, and key implementation barriers.

Most countries reported having national HBV and HCV guidelines aligned with international standards, although planned updates were inconsistent. Point of care PCR viral load testing was available in five countries, but coverage of test costs varied, and reflex PCR testing had been implemented in only four. First line antiviral regimens largely followed international guidance; however, medication prices and national coverage differed considerably, with out of pocket payment requirements persisting in several settings. All countries reported the use of non invasive tests for liver disease staging, though coverage for elastography remains limited. Service delivery has become increasingly decentralized, with HBV and HCV testing and care available across multiple levels of the health system and integrated into primary care and HIV clinics in most countries. Nevertheless, provision of HCV treatment in harm reduction settings remains rare, limiting access for populations at highest risk.

Overall, the findings indicate strong political commitment but highlight persistent gaps in testing access, treatment affordability, and service delivery models. Addressing these gaps through tailored country specific strategies, expanded financial protection, and strengthened integration of services will be essential to accelerate progress toward the 2030 elimination targets.

## Background

The World Health Organization (WHO) European Region is estimated to have 10,600,000 people living with hepatitis B infection and 7,700,000 people living with hepatitis C infection as of the end of 2022. However, only 15.7% of hepatitis B and 29% of hepatitis C cases were diagnosed, and only 1.9% of hepatitis B and 9.0% of hepatitis C cases received treatment (1). The number of new hepatitis B infections in 2022 was estimated at 18,000, and hepatitis C infections at 126,000 (1). WHO European Regional Action Plan for HIV, viral hepatitis and sexually transmitted infections has an ambitious target of reducing the incidence of hepatitis B to 2,200 and of hepatitis C to 25,000 by 2030 (2). To identify country-level implementation and barriers in achieving the elimination of hepatitis by 2030, WHO Regional Office for Europe conducted a survey on current policies and practices for testing and treatment of chronic hepatitis B and C infection in selected high-burden countries.

### WHO survey on national policy and practice for hepatitis B and C infections

The reporting form was sent through the WHO Country Offices to the following nine countries between November 2024 and April 2025: Armenia, Azerbaijan, Belarus, Kazakhstan, Kyrgyzstan, the Republic of Moldova, Tajikistan, Turkmenistan, and Uzbekistan. The selection of countries was based on 1) Burden of hepatitis B infections and hepatitis C infections (see Supplementary Files Figures S1 and S2), 2) Country’s capacity in control of hepatitis B and C infections, and 3) availability and operational capacity of WHO Country Offices. National focal points were asked to complete the form in English or Russian. The survey included questions on national guidelines, testing and treatment policies, service delivery and country-specific challenges in scaling up hepatitis B virus (HBV) and hepatitis C virus (HCV) testing and treatment.

### Survey results

We received completed forms from eight countries (response rate 89%): Armenia (ARM), Azerbaijan (AZE), Belarus (BLR), Kazakhstan (KAZ), Kyrgyzstan (KGZ), the Republic of Moldova (MDA), Turkmenistan (TKM) and Uzbekistan (UZB).

#### National guidelines

Seven countries reported having a national guideline for HBV and eight for HCV, with publication years ranging from 2012 to 2024 (Table 1). All available national guidelines referred to at least one international standard guideline among WHO, the European Association for the Study of the Liver (EASL), or the American Association for the Study of Liver Diseases (AASLD). Of the 15 available HBV or HCV national guidelines, updates are planned within five years of publication for six (40%).

**Table 1.** National guideline for testing and treatment of chronic hepatitis B and C infections.

|  | Armenia |  | Azerbaijan |  | Belarus |  | Kazakhstan |  | Kyrgyzstan |  | Republic of Moldova |  | Turkmenistan |  | Uzbekistan |  |
| --- | --- | --- | --- | --- | --- | --- | --- | --- | --- | --- | --- | --- | --- | --- | --- | --- |
|  | HBV | HCV | HBV | HCV | HBV | HCV | HBV | HCV | HBV | HCV | HBV | HCV | HBV | HCV | HBV | HCV |
| National guideline | Yes | Yes | N/A | Yes | Yes | Yes | Yes | Yes | Yes | Yes | Yes | Yes | Yes | Yes | Yes | Yes |
| Publication year | 2020 |  |  | 2021 | 2019 | 2019 | 2019 | 2020 | 2024 |  | 2021 | 2024 | 2012 | 2024 | 2024 |  |
| Referred international guideline | WHO, EASL |  |  | WHO, EASL, AASLD | EASL | EASL | WHO, EASL, AASLD | EASL, AASLD | WHO, EASL, AASLD |  | WHO, EASL, AASLD |  | WHO, EASL |  | WHO, EASL |  |
| Planned update year | 2025 |  |  | Unknown | 2025-26 | 2025-26 | 2025-26 | 2025-26 | TBD |  | 2025 | TBD | 2024 | Unknown | 2026 |  |

AASLD: American Association for the Study of Liver Diseases, EASL: European Association for the Study of the Liver, N/A: not available, U/K: unknown, TBD: to be decided, WHO: World Health Organization.

#### Point-of-care and reflex PCR testing availability

The availability of point-of-care (POC) polymerase chain reaction (PCR) testing for HBV and HCV was assessed, along with associated costs and whether these were covered by national programs (Table 2). HBV and HCV POC PCR were available in five countries: Belarus, Kazakhstan, Kyrgyzstan, Turkmenistan and Uzbekistan. Two countries with available data reported similar costs of $8–$9 USD. The POC PCR cost was fully covered in two countries, partially covered in one, and only HCV testing was covered in one. One country did not report coverage. Additionally, the survey asked about the implementation of reflex PCR testing; only four countries had implemented reflex testing for HBV and/or HCV.

**Table 2.**
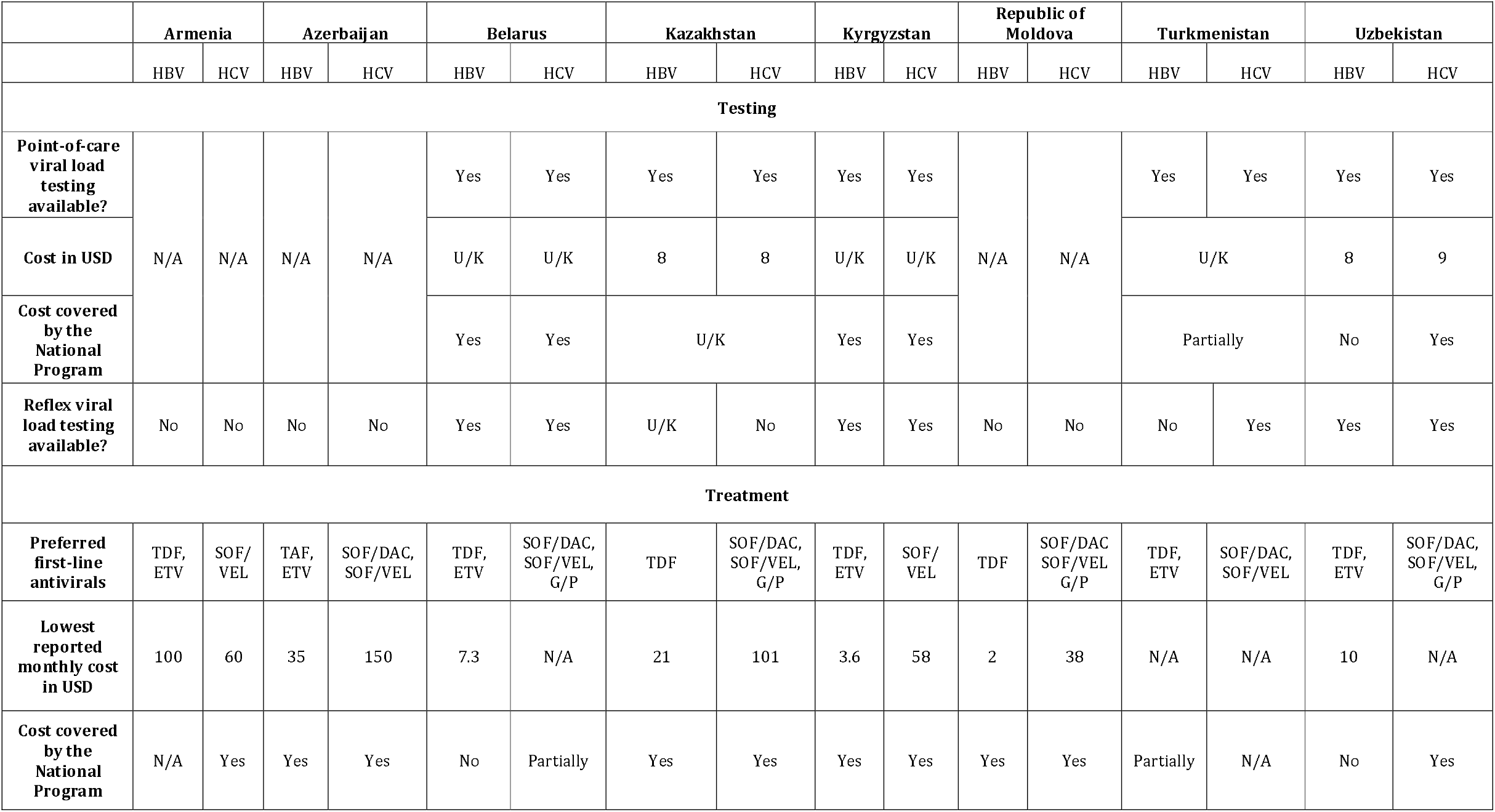
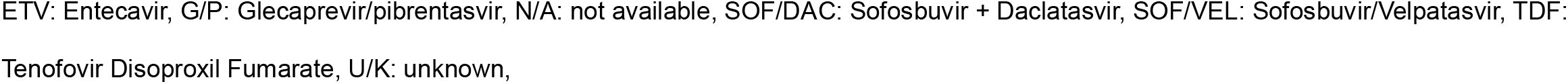
Testing and treatment for chronic hepatitis b and c infections.

|  | Armenia |  | Azerbaijan |  | Belarus |  | Kazakhstan |  | Kyrgyzstan |  | Republic of Moldova |  | Turkmenistan |  | Uzbekistan |  |
| --- | --- | --- | --- | --- | --- | --- | --- | --- | --- | --- | --- | --- | --- | --- | --- | --- |
|  | HBV | HCV | HBV | HCV | HBV | HCV | HBV | HCV | HBV | HCV | HBV | HCV | HBV | HCV | HBV | HCV |
| **Testing** |  |  |  |  |  |  |  |  |  |  |  |  |  |  |  |  |
| **Point-of-care viral load testing available?** | N/A | N/A | N/A | N/A | Yes | Yes | Yes | Yes | Yes | Yes | N/A | N/A | Yes | Yes | Yes | Yes |
| **Cost in USD** |  |  |  |  | U/K | U/K | 8 | 8 | U/K | U/K |  |  | U/K |  | 8 | 9 |
| **Cost covered by the National Program** |  |  |  |  | Yes | Yes | U/K |  | Yes | Yes |  |  | Partially |  | No | Yes |
| **Reflex viral load testing available?** | No | No | No | No | Yes | Yes | U/K | No | Yes | Yes | No | No | No | Yes | Yes | Yes |
| **Treatment** |  |  |  |  |  |  |  |  |  |  |  |  |  |  |  |  |
| **Preferred first-line antivirals** | TDF, ETV | SOF/VEL | TAF, ETV | SOF/DAC, SOF/VEL | TDF, ETV | SOF/DAC, SOF/VEL, G/P | TDF | SOF/DAC, SOF/VEL, G/P | TDF, ETV | SOF/VEL | TDF | SOF/DAC, SOF/VEL, G/P | TDF, ETV | SOF/DAC, SOF/VEL | TDF, ETV | SOF/DAC, SOF/VEL, G/P |
| **Lowest reported monthly cost in USD** | 100 | 60 | 35 | 150 | 7.3 | N/A | 21 | 101 | 3.6 | 58 | 2 | 38 | N/A | N/A | 10 | N/A |
| **Cost covered by the National Program** | N/A | Yes | Yes | Yes | No | Partially | Yes | Yes | Yes | Yes | Yes | Yes | Partially | N/A | No | Yes |

ETV: Entecavir, G/P: Glecaprevir/pibrentasvir, N/A: not available, SOF/DAC: Sofosbuvir + Daclatasvir, SOF/VEL: Sofosbuvir/Velpatasvir, TDF:
Tenofovir Disoproxil Fumarate, U/K: unknown,

#### Preferred first-line antivirals and their cost

Countries reported their preferred first-line antivirals and monthly costs (Table 2). For HBV, five countries recommend Tenofovir Disoproxil Fumarate (TDF) or Entecavir (ETV), and three recommend TDF alone. For HCV, all countries recommended Sofosbuvir/Velpatasvir (SOF/VEL), six included Sofosbuvir + Daclatasvir (SOF/DAC), and four recommended Glecaprevir/Pibrentasvir (G/P). Monthly costs varied widely: for HBV from $2 to $100 USD, and for HCV from $38 to $300 USD. The cost of antiviral medication is fully covered in four countries (50%). In two countries, only HCV antivirals are covered; HBV antivirals cost $7.3–$10 per month. One country reported partial HBV coverage, and one reported partial HCV coverage (costs unknown).

#### Non-invasive test for liver disease staging

Countries reported tools used for liver disease staging (Figure 1). All reported using non-invasive tests; liver elastography and the Aspartate Aminotransferase-to-Platelet Ratio Index (APRI) were most common. A mixed approach was frequently used. Cost coverage varied: three countries fully covered non-invasive tests, while four covered only the blood tests needed for APRI, not elastography. One country reported partial coverage (details unavailable).

**Figure 1.**
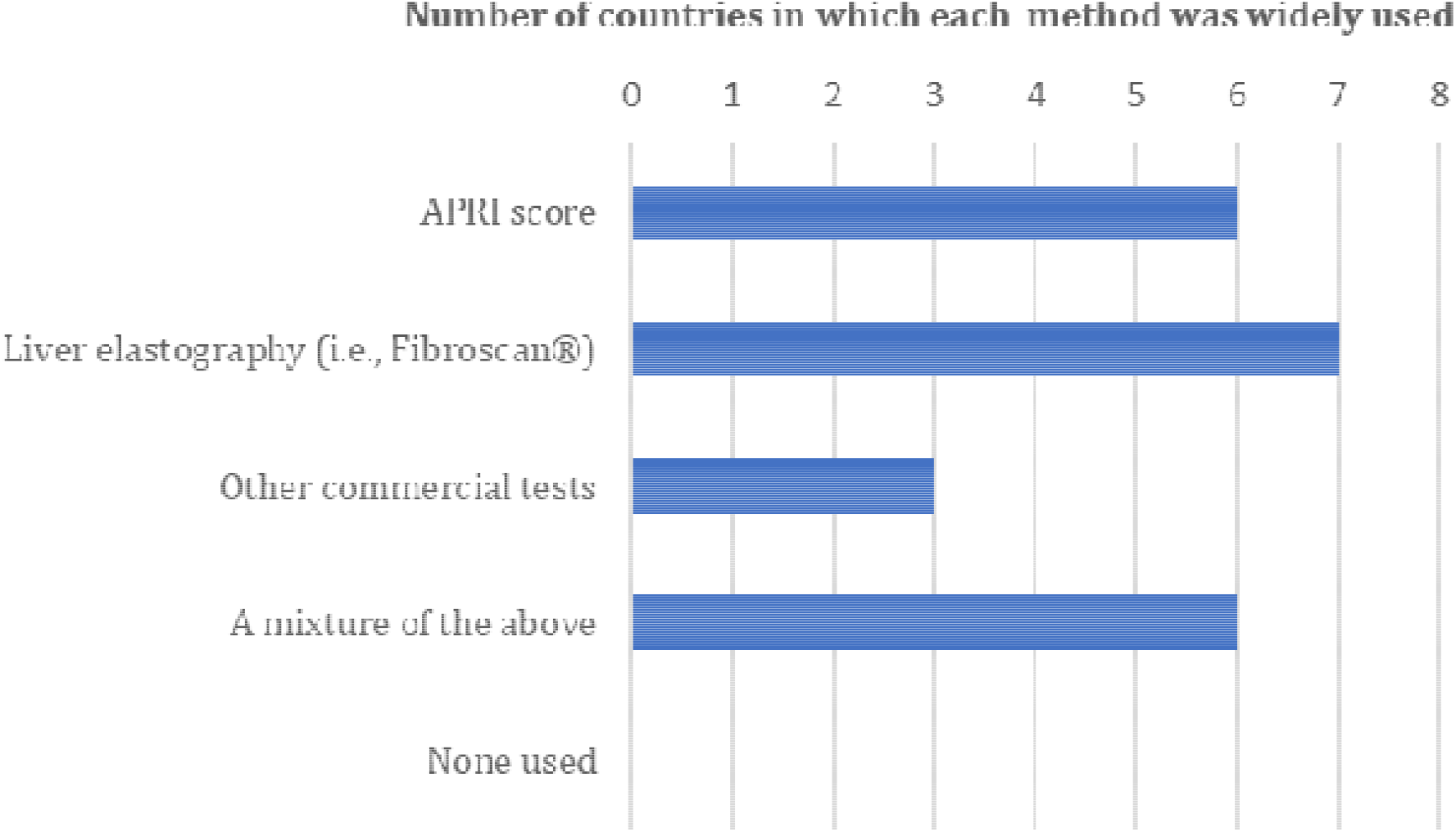
Non-invasive tests implementation in eight high-burden countries in the WHO European Region. APRI: Aspartate Aminotransferase-to-Platelet Ratio Index The number of countries in which each non-invasive test is widely used is shown. Other commercial tests include Fibrotest®/Fibrosure®, Fibrometer™.

#### Care and treatment service delivery sites

Countries described current service delivery sites for HBV and HCV care (Figure 2). Care was available across multiple hospital levels, reflecting effective decentralization. In six of eight countries, people could access HBV and HCV care at primary health care sites and HIV clinics, showing integration into existing services. However, only one country provided HCV treatment in harm-reduction sites, and none provided HBV care there. Three countries allowed non-specialist doctors or nurses to provide HBV and/or HCV care.

**Figure 2.**
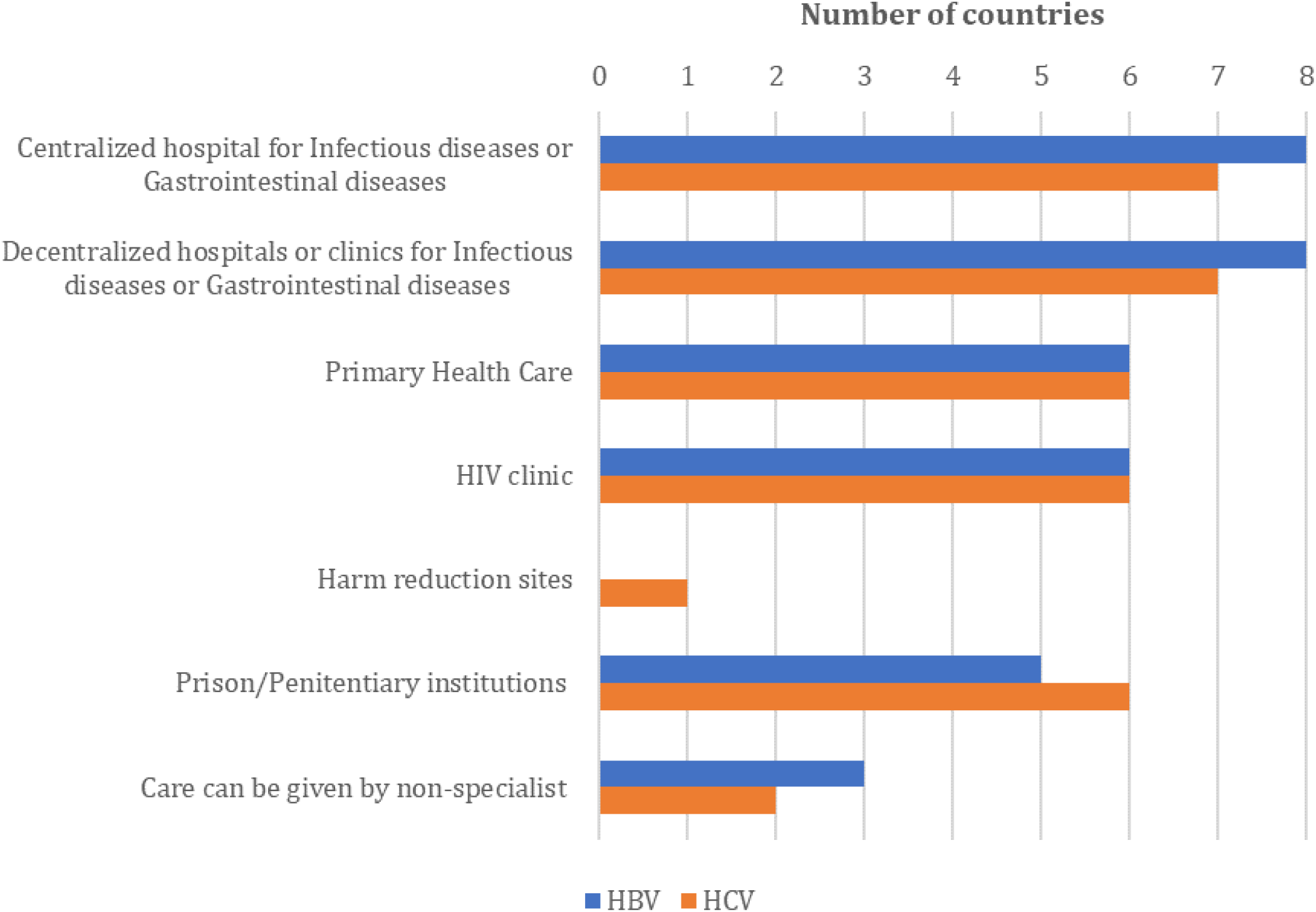
Service delivery sites and availability of services by non-specialists. The number of countries in which HBV and HCV care and treatment are provided in each facility, and by non-specialists if necessary.

#### Country-specific key challenges in scaling up HBV and HCV testing and treatment

Countries identified a range of challenges, including: absence of a national HBV guideline (AZE), lack of universal screening (KAZ), unstable supply of testing reagents (TKM), inadequate linkage to care (MDA), limited access to treatment (ARM), disrupted antiviral supply chains (AZE), out-of-pocket payments (ARM, UZB), shortage of skilled human resources (AZE), insufficient healthcare infrastructure (MDA), limited awareness and education (MDA), difficulty reaching high-risk populations (MDA), stigma (MDA) and gaps in preventing vertical transmission of HBV (MDA).

### Identified progress and gaps in the testing and treatment of chronic hepatitis B and C infection

The reporting countries demonstrated strong commitments to HBV and HCV control through national guidelines that follow international standards and expressed intentions to update these guidelines in the coming years. Countries that do not have a guideline or have a guideline published more than five years ago are recommended to consider generating or revising their guidelines.

The availability of PoC and reflex PCR testing varied between countries. One of the biggest challenges along the viral hepatitis care cascade has been limited access to viral load testing in resource-constrained settings because of high costs and requirements for specialised laboratory infrastructure, trained personnel and a sample transport system (3)(4). WHO recommends PoC HBV DNA and PoC HCV RNA testing as alternative approaches to laboratory-based platforms to enhance access to and uptake of testing and treatment for HBV (5) and HCV (6). Also, reflex viral load testing in those with a positive HBsAg test result for HBV DNA test and a positive HCV antibody test result for HCV RNA has been added in the WHO recommendations as additional strategies to promote linkage to care and treatment.

Preferred first-line regimen in high-burden countries is aligned with international standard guidelines. While most countries reported full coverage of the cost of antivirals by their national program, there are a few countries where out-of-pocket payment is required for HBV or HCV treatment. Financial barriers often impede access to care, and removing them is essential to realise equitable access to treatment regardless of the patient’s economic status (7).

Non-invasive tests for liver disease staging are widely implemented in the reporting countries. The cost of testing is fully or partially covered by the national program in all countries, covering at least the cost of the APRI score. In the latest WHO guideline, non-invasive tests are recommended for assessing treatment eligibility, monitoring treatment response, and/or disease progression (5)(6). Ensuring access to non-invasive tests without a financial burden is crucial to expand the testing and treatment coverage of people living with HBV and/or HCV, especially in resource-limited settings (8). Therefore, countries should make efforts to remove financial barriers to improve access to diagnosis.

While HBV and HCV care and treatment are delivered in various levels of facilities, only one country reported the availability of HCV care and treatment in harm reduction sites. Providing HCV care and treatment in harm reduction sites is included in the WHO recommendations (6). Countries should consider expanding service delivery sites further to reach people at high risk of acquiring HCV more effectively.

Although this survey does not cover all countries in the WHO European Region due to coordination difficulties, the collected data revealed shared challenges in the selected high-burden countries, including inadequate cost coverage by national programs and the need for further expansion of testing options and service delivery. At the same time, a wide variety of the mentioned top challenges reflects the need for tailored approaches for each country.

## Data Availability

All available data collected in this study can be shared upon reasonable request after acquiring approval from the respective countries.

## Declarations

### Ethics approval and consent to participate

Ethical approval was not necessary for this study because no individual-level data were collected.

### Authors’ contributions

MO led the data collection process, wrote the manuscript, and generated tables and figures.

MN and SB contributed to the data collection, reviewed the manuscript and agreed on its final version.

## Acknowledgement

The authors would like to thank the national hepatitis program managers who participated in the survey and colleagues in the WHO country offices for their coordination.

## National hepatitis programmes

Armenia: Ministry of Health (MOH) of the Republic of Armenia, National Center for Infectious Diseases of the MOH Armenia, and National Center for Diseases Prevention and Control of the MOH Armenia

Azerbaijan: Ibrahim Rufullayev and Sugra Azimova (Administration of Regional Medical Divisions (TABIB))

Belarus: Dzmitry Danilau (Belarusian State Medical University) and Iryna Hlinskaya (Republican Center for Hygiene, Epidemiology and Public Health)

Kazakhstan: Kulpash Sagyndykovna Kaliaskarova, Gulnara Utepergenovna Kulkaeva, and Gulnara Yedilovna Sarsenbaeva (Ministry of Health of the Republic of Kazakhstan), Alma Serikpaevna Aubakirova (National Scientific Oncology Center)

Kyrgyzstan: Mambetisaeva Anara (Public Health Department, Ministry of Health), Chokmorova Umut (the Republican center for control of the blood borne viral hepatitis and HIV, Ministry of Health), and Kenzhekarieva Aidana (the Republican center for control of the blood borne viral hepatitis and HIV, Ministry of Health)

Republic of Moldova: Lucia Pirtina (Clinical Hospital for Infectious Diseases “Toma Ciorba”) and Lilia Cojuhari (State University of Medicine and Pharmacy “Nicolae Testemitanu”)

Turkmenistan: Aman Shukurov, Batyr Tajiyev and Jumadurdy Annanurov (Turkmen State Medical University named after M.O. Karryev)

Uzbekistan: Ismoilov Umed (Research Institute of Virology), Abdurakhimova Dilnoza (Republican Research Institute of Virology), and Joldasova Elizaveta (Republican Research Institute of Virology)

### WHO Country Offices

Gayane GHUKASYAN (Armenia), Sevinj HASANOVA (Azerbaijan), Oleg DOUBOVIK (Belarus), Stela GHEORGHITA (Kazakhstan), Elnura DUISHENALIEVA (Kyrgyzstan), Alexandru VOLOC (Republic of Moldova), Leyli SHAMYRADOVA (Turkmenistan), Jamshid GADOEV (Uzbekistan).

The views and opinions expressed in this paper are those of the authors and not necessarily the views, decisions or policies of the World Health Organization (WHO).

## Patient and Public Involvement

It was not appropriate or possible to involve patients or the public in the design, conduct, reporting, or dissemination plans of our research

