## Supplementary FIle for "CURRENT POLICIES AND PRACTICES FOR TESTING AND TREATMENT OF CHRONIC HEPATITIS B AND C INFECTION IN HIGH-BURDEN COUNTRIES IN THE WHO EUROPEAN REGION"

### Supplementary Files

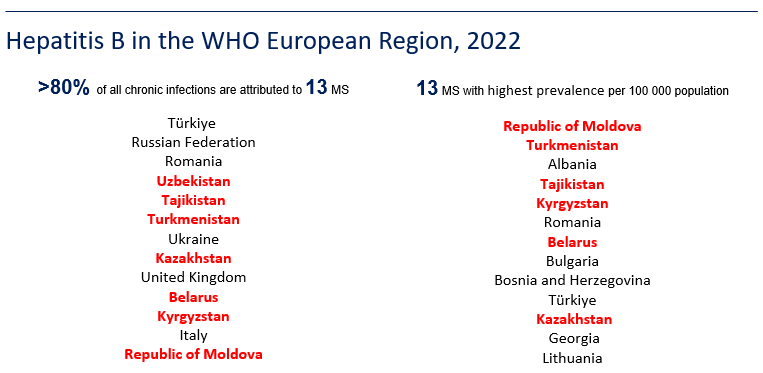

**Figure S1. Hepatitis B burden in the WHO European Region, 2022.**

Countries included in this study are highlighted in red.

Data based on WHO modelled estimates, 2022, collected for the World Hepatitis Report 2024.

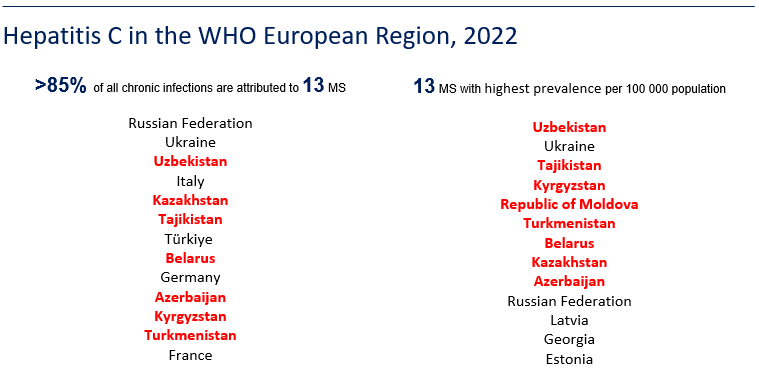

**Figure S2. Hepatitis C burden in the WHO European Region, 2022.**

Countries included in this study are highlighted in red.

Data based on WHO modelled estimates, 2022, collected for the World Hepatitis Report 2024.
